# Prognostic value and peripheral immunologic correlates of early FDG PET response imaging in a phase II trial of risk-adaptive chemoradiation for unresectable non-small cell lung cancer

**DOI:** 10.1101/2021.02.22.21251919

**Authors:** Stephen R. Bowen, Daniel S. Hippe, Hannah M. Thomas, Balukrishna Sasidharan, Paul D. Lampe, Christina S. Baik, Keith D. Eaton, Sylvia Lee, Renato G. Martins, Rafael Santana-Davila, Delphine Chen, Paul E. Kinahan, Robert S. Miyaoka, Hubert J. Vesselle, A. McGarry Houghton, Ramesh Rengan, Jing Zeng

**Author notes:** **Corresponding author:** Jing Zeng, MD, Associate Professor, Department of Radiation Oncology, University of Washington School of Medicine, 1959 NE Pacific St, Box 356043, Seattle, WA 98195. **Financial Support:** NIH/NCI R01CA204301.

## Abstract

**Introduction:** We hypothesized that FDG PET imaging during chemoradiation for unresectable non-small cell lung cancer (NSCLC) is prognostic for survival, and that tumor PET response is correlated with peripheral T-cell function.

**Methods:** 45 patients with AJCCv7 stage IIB-IIIB NSCLC enrolled on the phase II FLARE-RT trial and received platinum-doublet chemotherapy concurrent with 6 weeks of radiation (NCT02773238). FDG PET imaging was performed prior to treatment start and after 24 Gy of radiation (week 3). PET response status was prospectively defined by multifactorial radiologic interpretation. PET responders received 60 Gy in 30 fractions, while non-responders received concomitant boosts to 74 Gy in 30 fractions. Peripheral blood was drawn synchronously with PET imaging, from which germline DNA sequencing, T-cell receptor (TCR) sequencing, and plasma cytokines analysis were performed.

**Results:** Median follow-up was 18.8 months, 1-year overall survival (OS) 82%, 1-year progression-free survival (PFS) 53%, and 1-year locoregional control (LRC) 88%. Higher mid-treatment PET total lesion glycolysis was detrimental to OS (1-yr 87% vs. 63%, p<0.001), PFS (1-yr 60% vs. 26%, p=0.044) and LRC (1-yr 94% vs. 65%, p=0.012), even after adjustment for clinical/treatment factors. Higher PD-L1 tumor proportion score (TPS) was correlated with PET response (p=0.017): 6/6 patients with high PD-L1 (TPS>50%) were classified as PET responders, while 4/5 patients classified as PET non-responders had negative PD-L1 (TPS<1%). Higher TCR richness and clone distribution slope was associated with improved OS (p=0.018-0.035); clone distribution slope was correlated with PET response (p=0.031). Germline DNA alterations in immunologic pathways had an outsized effect on OS and PET response; of the top 30 SNPs ranked by association with PET response status (p<0.016), a plurality (13/30) came from immunologic pathways.

**Conclusions:** Mid-chemoradiation PET imaging is prognostic for survival; PET response may be linked to tumor and peripheral T-cell biomarkers.

## Introduction

The current standard of care for unresectable locally advanced non-small cell lung cancer (LA-NSCLC) is concurrent chemoradiation plus a year of adjuvant durvalumab, a programmed death-ligand 1 (PD-L1) immune checkpoint inhibitor, which improved survival over chemoradiation alone^1,2^. However, patient outcomes remain suboptimal even for well-selected clinical trial eligible patients, with an overall survival at 2 years of 66.3%^1^. Clinical trials seek to improve outcomes through intensification of therapy, both in terms of additional immune modulating therapy as well as localized radiation dose escalation. Beyond durvalumab, other PD-1/PD-L1 pathway checkpoint inhibitors are also being tested as consolidation therapy after chemoradiation, concurrent with chemoradiation, and neoadjuvant to chemoradiation^3^. As treatment intensifies, patient tolerance is increasingly challenged, with 29.1% of patients experiencing serious adverse events in the PACIFIC trial and 15.4% of patients discontinuing treatment due to adverse events. Although durvalumab improved 2-year overall survival (OS) from 55.6% to 66.3% in the PACIFIC trial, only a minority of the patients that received durvalumab derived a survival benefit from the additional treatment. A prognostic test that could predict patient outcome would help better select patients who need additional therapies, and spare toxicity in patients who do not.

FDG PET imaging during chemoradiation for NSCLC has been correlated with treatment response and survival^4-8^. Van Elmpt *et al*. found that for patients whose mean tumor standardized uptake values (SUV) decreased by >15% on mid-treatment PET, 2-year OS was 92% compared to 33% for patients with a decrease in mean SUV <15%^9^. Tumor FDG PET avidity has also been correlated with PD-L1 expression, with multiple clinical series suggesting SUVmax may be associated with PD-L1 positivity, and that SUVmax could be a potential predictive marker of response to anti-PD-1 therapy in NSCLC patients^10-12^.

In addition to predicting for survival, FDG PET avidity may also be predictive for local recurrences^13,14,^ both on the pre-chemoradiation scan and the mid-chemoradiation scan. Decades of trials to overcome local recurrence risk by radiation treatment intensification in unresectable NSCLC have yielded limited success. Results of RTOG 0617 showing inferior outcomes with 74 Gy versus 60 Gy during concurrent chemoradiation was against expectations and reset the standard of care radiation dose to around 60 Gy^15^. However, local control remains a problem in unresectable NSCLC with locoregional failures of around 20-30% by 2-years and rising to 50% by 3-5 years^1,16^,17. There are multiple theories regarding the lack of benefit with 74 Gy versus 60 Gy in RTOG 0617, including higher dose to normal tissues being detrimental (esophagitis grade and heart dose were both highly correlated with survival)^15^. Since uniform dose escalation in an unselected group of patients was detrimental to survival, multiple clinical trials are underway to test other strategies for dose escalation, to improve cancer control and survival. Some trials have tested giving all patients a radiation boost based on pre-treatment PET scans, targeting the most FDG-avid regions with a higher dose of radiation^18 19^, whereas other trials dose-escalate all patients based on mid-treatment PET^20^. Since some patients have durable cancer control after 60 Gy of radiation, not all patients may benefit from dose escalation. We designed a phase II trial FLARE-RT (NCT02773238) where select patients undergo dose escalation based on mid-treatment response as assessed by PET, and only non-responders undergo radiation dose escalation (74 Gy) for the second half of chemoradiation^21^.

We hypothesized that early FDG PET response imaging during chemoradiation has prognostic value in patients treated on the FLARE-RT trial, and that a robust tumor PET response is linked to peripheral T-cell function, given the clinical and preclinical data that a robust tumor response to radiation requires an intact immune system, including the presence of CD8+ T-cells^22,23.^ The purpose of this investigation was to (i) validate the prognostic value of early FDG PET response imaging, (ii) evaluate peripheral immunologic correlates of PET imaging, and (iii) identify complementary markers of tumor biology (PET response) and disease burden (PET total lesion glycolysis). Identification of biomarkers of early treatment response/resistance can support clinical management decisions, and inform the design of next-generation biomarker-guided and risk-adaptive clinical trials to improve outcomes in patients with LA-NSCLC.

## Materials and Methods

### Clinical Trial Protocol

The FLARE-RT phase II trial (NCT02773238) schema is shown in Figure 1. This research was IRB approved and conducted in accord with the ethical standards of our institution. Patients with histopathologically confirmed unresectable AJCCv7 stage IIB-IIIB non-small lung cancer and ECOG performance status 0-1 were enrolled from 2016-2020. Baseline FDG PET imaging was followed by initiation of definitive chemoradiation planned for 60 Gy in 30 fractions. Chemotherapy consisted of a platinum doublet per physician choice (Table 1) and started at the same time as radiation. Consolidation immunotherapy was allowed post-chemoradiation once it became standard of care. During the third week of chemoradiation (after 24 Gy delivered), FDG PET imaging was performed to assess early treatment response. Patients prospectively classified as responders continued to receive a total dose of 60 Gy in 30 fractions, while those classified as non-responders received a concomitant dose boost over the final 15 fractions to a total dose of 74 Gy in 30 fractions, as previously described^21^. The precision dose boost was spatially localized and redistributed to conform to residual intratumoral FDG avidity. Peripheral blood draws were completed synchronously with PET imaging at baseline, week 3 mid-treatment, and 3-month post-treatment time points. The primary trial endpoint was 2-year OS, while secondary endpoints included 1-year PFS, 1-year LRC, and pulmonary toxicity (CTCAE v4 grade 2+ pneumonitis).

**Figure 1.**
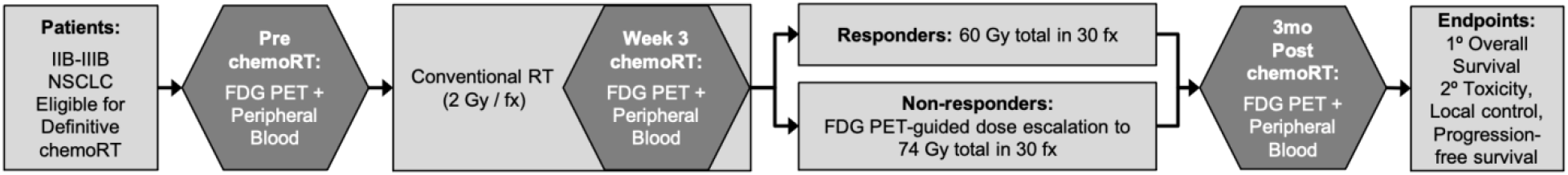
FLARE-RT phase II trial schema of risk-adaptive chemoradiation for patients with unresectable locally advanced non-small cell lung cancer.

**Table 1.** Patient characteristics (n=45).

| Characteristic | N (%) or median (range) |
| --- | --- |
| Age (years) | 63 (34-78) |
| Gender |  |
| Female | 25 (63%) |
| Male | 20 (37%) |
| Histology |  |
| Adenocarcinoma | 29 (64%) |
| Squamous Cell Carcinoma | 14 (31%) |
| Other | 2 (4%) |
| Chemotherapy |  |
| Carboplatin-Paclitaxel | 25 (56%) |
| Cisplatin-Etoposide | 11 (24%) |
| Other Platinum Doublet | 9 (20%) |
| Stage (AJCC v7) |  |
| IIB | 2 (4%) |
| IIIA | 23 (51%) |
| IIIB | 15 (33%) |
| N2 Recurrence | 5 (11%) |
| Radiation Therapy |  |
| Photon IMRT | 22 (49%) |
| Proton Beam Radiation | 23 (51%) |
| Consolidation Immune Checkpoint Inhibitor |  |
| Yes | 23 (51%) |
| No | 22 (49%) |
| PD-L1 Tumor Proportion Score |  |
| >50% | 6 (13%) |
| 1-49% | 7 (16%) |
| <1% | 7 (16%) |
| Unknown | 25 (56%) |
| Driver Mutation (EGFR/ALK/ROS1) |  |
| Yes | 9 (20%) |
| No | 19 (42%) |
| Unknown | 17 (38%) |
| Mid-PET FLARE Response |  |
| Responder | 29 (64%) |
| Non-responder | 16 (36%) |
| Mid-PET PERCIST 1.0 |  |
| Partial Metabolic Responder | 27 (60%) |
| Stable Metabolic Disease | 17 (38%) |
| Progressive Metabolic Disease | 1 (2%) |

### PET/CT Response Assessment & Residual Total Lesion Glycolysis

Week 3 PET response status was prospectively defined by multifactorial radiologic interpretation. Quantitative assessments were based on interval changes in standardized uptake value (SUV) metrics (SUVmax, SUVmean, SUVpeak), metabolic tumor volume (MTV), and total lesion glycolysis (TLG) using semi-automatic gradient-based segmentation of the primary tumor and adjacent involved lymph nodes. PET responders presented with greater than 20% decrease in at least one metric of FDG avidity (SUVmax, SUVmean, SUVpeak) and at least one metric of FDG volumetric extent (MTV, TLG), along with clinical radiographic evidence of interval response to treatment. Metabolic tumor volume contours were defined by a commercially validated segmentation algorithm (PET Edge, MIM Software, Cleveland, OH) that achieved improved inter-observer agreement compared to manual contouring and reduced sensitivity to image reconstruction compared to fixed threshold contouring^24,25.^ Prospective FLARE protocol-defined mid-chemoradiation PET response status showed substantial agreement to retrospective mid-chemoradiation PET response assessment by PERCIST 1.0^26,27^ (Kappa 0.72 [0.51-0.93]). Residual total lesion glycolysis on week 3 PET (TLGmidtx [g]) was calculated as the product of MTV [mL] and SUVmean [g/mL]. Given the skewed distribution of TLGmidtx across patients, we dichotomized the upper tail of the distribution from the bulk, which was achieved at the 80^th^ percentile corresponding to a threshold of TLGmidtx = 250 g. Dichotomized TLGmidtx defined patient risk strata in survival analyses.

### Peripheral Blood Genomic & Immunologic Assays

Sequencing of peripheral germline DNA was conducted using the Infinium Global Screening Array (Illumina Inc. San Diego, CA), which included sample QC, library prep+QC, cluster optimization, and sequencing steps. Sequencing libraries were prepared with a Covaris LE220 system and PerkinElmer Sciclone NGSx workstation. Library QC was ensured with two Agilent Bioanalyzer 2100, an Agilent 2200 TapeStation, a Life Technologies Qubit 2.0 fluorometer and an ABI StepOne Real-Time PCR system. Genotypes with GenCall scores > 0.15 were considered of sufficient quality for inclusion. Genome sequencing reports were filtered to include single nucleotide polymorphism (SNP) genotypes from a list of predefined relevant candidate genes (see Supplemental Table 1) belonging to different pathway families: 96 DNA repair genes^28,29,^ 53 immunology genes^30-32^, 38 oncology genes^33,34^, and 27 lung biology genes^35,36.^

Molecular T-cell receptor (TCR) beta chain CDR3 sequencing was carried out on the ImmunoSEQ platform (Adaptive Biotechnologies, Snohomish, WA) with a survey sampling depth of 120,000 T-cells. We characterized the immune repertoire through diversity metrics, which capture the number of unique T-cell receptors (richness) and the relative abundance of each receptor (clonality / evenness). Richness was averaged over the following measures due to high collinearity (Spearman r > 0.93): iChao1^37^, Efron Thisted Estimator^38^, and Daley Smith Estimator^39^. Clonality / evenness measures included the following: Pielou Evenness^40^, Simpson Clonality and Evenness^41^, and Clone Distribution Slope^42^. In general, higher richness and higher clone distribution slope indicate greater TCR diversity with the presence of more rare clones.

Exploratory single immune cell functional assays were performed for 4 patients on the IsoLight system using IsoCode Human Adaptive Immune chips (Isoplexis, Branford, CT), which can measure 25+ cytokine secretions on a single cell base of approximately 1000 individual cells per chip. We restricted our analysis to abundance of polyfunctional CD8+ T-cells, following stimulation and secretion of at least 2 effector cytokines (e.g. MIP1β, IFNγ). Lastly, detectable plasma concentrations from an immunologic panel of 43 cytokines were measured via a combination of the Luminex 200 microbead system (Luminex, Austin TX) and standard ELISA.

### Statistical Analysis

Overall survival (OS), progression-free survival (PFS), and distant metastatic-free (DM) survival rates were estimated by Kaplan-Meier. Locoregional control (LRC) and pneumonitis (PNM) rates were adjusted for distant progression and death as competing risks. Differences between risk strata were evaluated with log rank or Gray’s testing. Univariate and bivariate hazard ratios (HR) were estimated by Cox regression or Fine & Gray competing risk regression.

Unsupervised learning of peripheral germline DNA sequencing SNP genotypes was performed with hierarchical clustering and summarized with cumulative frequency distributions of rank ordered SNPs for each genetic pathway family (DNA repair, immunology, oncology, lung biology). Genotype associations to PET response status were characterized by logistic regression with Firth penalization for estimating odds ratios (OR). Genotype associations to OS and PFS were characterized by Cox regression with Firth penalization for estimating hazard ratios (HR). Genotype associations to TLGmidtx were characterized by Spearman rank correlation.

Associations between PD-L1 tumor proportion score (TPS) and PET response status were summarized by logistic regression with Firth penalization, Spearman rank correlation, and Fisher exact testing. PD-L1 TPS associations with TLGmidtx were estimated by Spearman rank correlation, while associations to histology, stage, and presence of driver mutation were estimated by Fisher exact testing.

TCR diversity metric trends over pre-treatment, mid-treatment, and post-treatment timepoints were estimated by Spearman rank correlation and pairwise changes by Wilcoxon signed rank testing. TCR diversity metric associations to OS and PFS were assessed by Permutation testing of Harrell’s c-index. TCR diversity metric associations to PET response class were assessed by groupwise Wilcoxon rank sum testing. TCR diversity metric associations to TLGmidtx were estimated by Spearman rank correlation.

Unsupervised learning of plasma cytokines was performed with hierarchical clustering and summarized with a 2-dimensional dendrogram heatmap of cytokine concentration z-scores with rugs for OS, PFS, PET response, and TLGmidtx classes. Associations of cytokines with PET response status were assessed by Fisher exact testing, associations with OS and PFS were estimated by Cox regression with Firth penalization, and associations with TLGmidtx were estimated by Spearman rank correlation. All statistical analyses and data visualization were carried out in OriginPro 2020b (OriginLab, Northampton, MA) and R version 4.0.3 (R Statistical Language, Vienna, Austria).

Throughout, two-sided tests were used with p < 0.05 considered statistically significant, without adjustment for the number of comparisons.

## Results

### Patient Characteristics

Patient characteristics are summarized in Table 1. Forty-nine patients were enrolled but 4 patients did not initiate any clinical trial therapy (3 were found to have metastatic disease on repeat baseline imaging, and 1 patient chose to have surgery instead). In the intention-to-treat population (n=45) with median age of 63 years (range 34-78 years), most patients enrolled on the FLARE-RT trial had unresectable, stage III, multi-station N2 positive NSCLC. Approximately half of patients received consolidation immune checkpoint inhibitor therapy when the standard of care changed mid-point during this trial (Table 1), and about half the patients were treated with proton beam radiation (Table 1) and half with photon intensity-modulated radiation therapy (IMRT).

### Clinical Outcomes

Figure 2 summarizes clinical outcomes of the FLARE-RT phase II trial from the time of consent. The median time between consent and start of chemoradiation was 11 days (range: 0 - 24 days). After a median follow-up of 18.8 months (range 3.0-49.9 months) in the intention-to-treat (ITT) population (n=45), 1-year OS was 82%, 1-year PFS was 53%, 1-year LRC was 88%, 1-year DM was 61%, and 6-month pneumonitis (PNM) cumulative incidence was 25%. Two of 45 patients terminated chemoradiation early due to progressive disease during chemoradiation, and switched to systemic therapy alone, but were included in the ITT analysis. Patients who received durvalumab immune checkpoint inhibitor therapy had numerically higher cumulative incidence of grade 2+ pneumonitis versus those who did not (1-yr PNM 52% vs. 26%, Gray p=0.20). Receipt of consolidation durvalumab according to the PACIFIC regimen resulted in numerically higher PFS but did not reach statistical significance in our trial cohort (median PFS 19.4 months vs. 10.9 months, log rank p=0.45).

**Figure 2.**
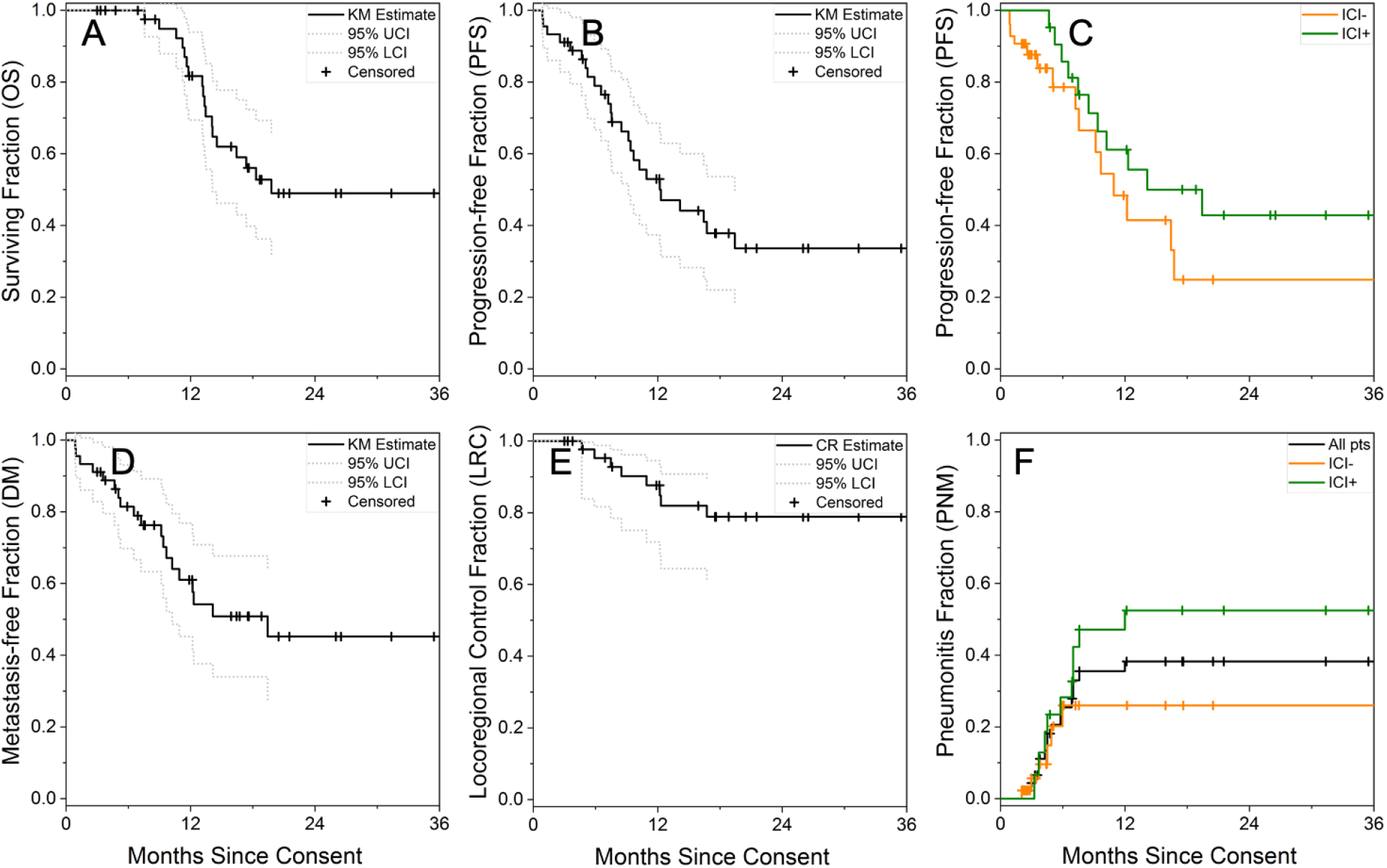
FLARE-RT clinical trial outcomes: overall survival (OS, A), progression-free survival (PFS, B), PFS stratified by receipt of consolidation immune checkpoint inhibitor therapy (ICI, C), distant metastatic-free survival (DM, D), locoregional control (LRC, E), CTCAE v4 grade 2 or higher pneumonitis (PNM, F). KM: Kaplan-Meier, CR: competing risk (distant progression / death).

### Week 3 Mid-treatment PET Risk Stratification

Figure 3A-C shows that residual total lesion glycolysis (TLGmidtx) on week 3 PET, dichotomized at the 80^th^ percentile (250 g), was significantly associated with a detriment in OS (log rank p < 0.001), PFS (log rank p = 0.044), and LRC (Gray p = 0.012). In the setting of FLARE-RT trial protocol PET response-adaptive dose escalation, PET response status was not associated with outcomes (p > 0.25). However, Figure 3D captures a highly significant interaction between PET response status and TLGmidtx that modulated PFS (log rank p < 0.00001), in which the highest risk of rapid disease progression was in PET non-responders with TLGmidtx > 250 g. Of the 9 patients with TLGmidtx > 250 g, 6/9 were classified as PET responders and received standard chemoradiation without PET-guided dose escalation. Forest plots in Figure 3E-F summarize the associations of TLGmidtx with risk of death or risk of disease progression, which were maintained after bivariate adjustment with clinical and treatment factors, including radiation target volume (PTVpretx) and PET response status.

**Figure 3.**
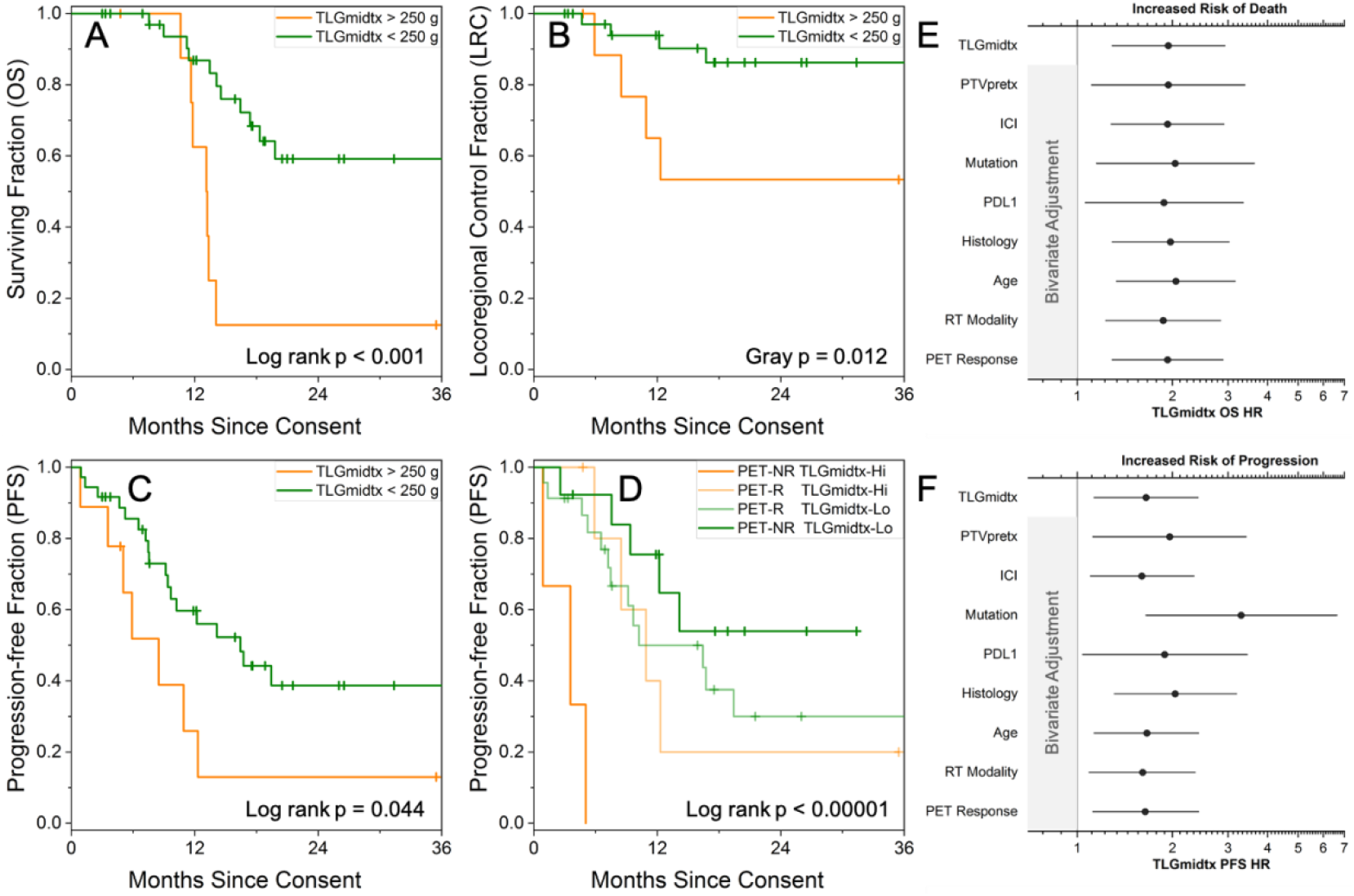
Kaplan-Meier overall survival (OS), competing risk-adjusted locoregional control (LRC), and Kaplan-Meier progression-free survival (PFS) stratified by week 3 mid-treatment FDG PET total lesion glycolysis (TLGmidtx) (A-C), along with interaction of total lesion glycolysis with PET response status for PFS (D). Forest plots of total lesion glycolysis OS and PFS univariate and bivariate hazard ratios, adjusted for individual clinical and treatment factors (E-F). PET-NR: PET non-responder; PET-R: PET responder.

### Peripheral Germline DNA Sequencing

Figure 4 depicts the cumulative frequency of SNP genotypes of different genetic pathway families (DNA repair, immunology, lung biology, oncology) in 19 patients rank ordered by their effect size on unsupervised hierarchical cluster membership (A), risk of death (B), PET response class membership (C), and residual total lesion glycolysis correlation (D). A larger frequency of SNP gene alterations in the immunology pathways belong to the main hierarchical cluster and have significant association to overall survival (Figure 4A-B), relative to other pathway families which reside closer to the diagonal reference line of equal frequency contribution. Differences in gene alteration frequency between FLARE-RT PET non-responder and responder subgroups were observed across several pathway families, particularly immunology and oncology (Figure 4C). By contrast, there was no preferential correlation of specific pathway families with total lesion glycolysis, which all have cumulative frequency distributions that closely follow the diagonal reference line (Figure 4D). Of the top 30 SNPs ranked by association with PET response status (p < 0.016), a plurality (13/30) came from immunologic pathways, while none of these same SNPs were associated with total lesion glycolysis (p > 0.11). Immunologic pathway genetic differences in JAK1 associated with PET response status and PFS (p=0.033-0.040), while differences in STAT1 (p=0.001-0.030) and IFNγ (p=0.002-0.060) associated with PET response status and hierarchical cluster membership. Other associations with PET response status included germline genetic alterations in VEGFC (p=0.002), ALK (p=0.018) and EGFR (p=0.047). Associations with OS included immuno-oncogene alterations in ALK (p=0.012), PIK3CA (p=0.013) and TNF (p=0.041). None of these gene alterations were associated with total lesion glycolysis (p>0.38). Without adjustments for multiple comparisons, our results are hypothesis generating rather than conclusive.

**Figure 4.**
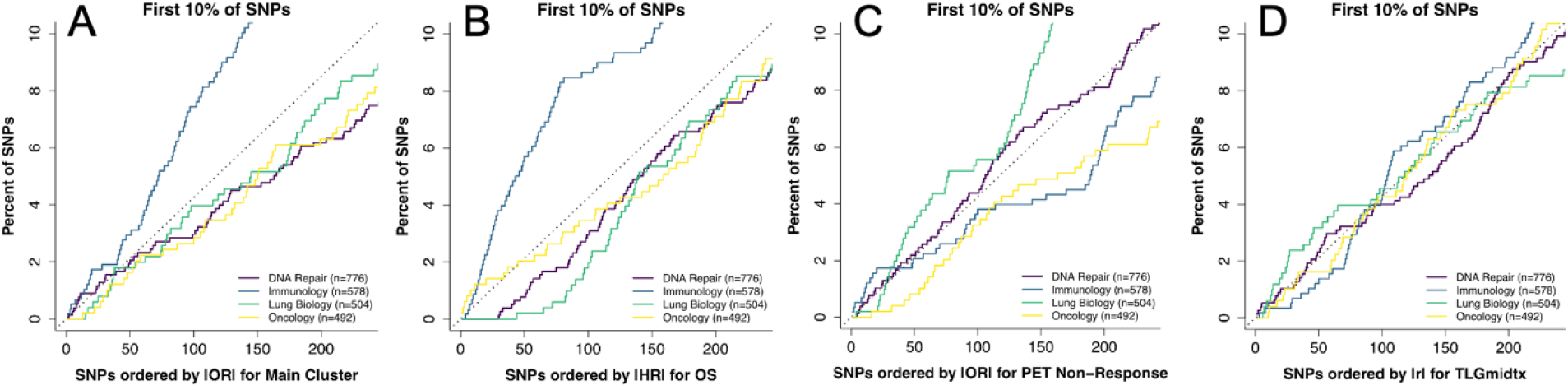
Peripheral DNA microarray cumulative frequency of single nucleotide polymorphism (SNP) gene alterations by pathways between unsupervised hierarchical clusters (A), differential risk of death (B), PET response groups (C), and PET total lesion glycolysis (TLGmidtx) (D). The diagonal reference line represents equal frequency contributions from all SNPs. The frequency of gene alterations in immunologic pathways has an outsized effect on risk of death and hierarchical cluster membership relative to other pathways. PET response status shows highly significant association to gene alterations across several pathways, while PET TLGmidtx correlations to gene alterations are more randomly distributed without linkages to specific pathways. OR: odds ratio, HR: hazard ratio, r: Spearman rank correlation.

### PET Response vs. Tumor PD-L1 Expression

PET response status was correlated with PD-L1 tumor proportion score (TPS) in 20 patients: 6/6 patients with high PD-L1 TPS (≥50%) were PET responders and 6/7 with moderate PD-L1 TPS (1-49%) were PET responders. By contrast, 4/5 patients classified as PET non-responders had negative PD-L1 TPS (<1%). The association between PET response and PD-L1 TPS was statistically significant as a trend across 3 expression levels (<1%, 1-49%, ≥50%, Firth p=0.017), rank correlation (Spearman r = 0.54, p=0.014) and categorical frequency (<1% vs. ≥1%, Fisher p=0.031). The association to PET response status was not confounded by availability of PD-L1 testing results: 15/20 were PET responders when PD-L1 was available, compared to 14/25 PET responders when unavailable (Fisher p=0.22). Tumor PD-L1 expression was neither correlated to mid-treatment total lesion glycolysis (Spearman r = 0.04, p=0.88) nor associated to histology, stage, or presence of a driver mutation (p>0.28).

### TCR Diversity Metrics & CD8+ T-cell Polyfunctionality

TCR sequencing was performed at pre-treatment (n=15), mid-treatment (n=15), and post-treatment (n=10) time points. No patients received immunotherapy pre-chemoradiation treatment or mid-treatment, and 10/10 patients were receiving immunotherapy (durvalumab) at the post-treatment time point. TCR average richness declined significantly across time points (Figure 5A, p=0.001). Pairwise changes were most significant between pre-treatment and post-treatment time points (Wilcoxon signed rank p=0.037). Higher pre-treatment TCR richness (Figure 5B, Permutation p=0.018), higher pre-treatment clone distribution slope (Figure 5D, Permutation p=0.035), and smaller decline in clone distribution slope (Permutation p=0.050) were associated with improved OS. Pre-treatment TCR clone distribution slope, pre- and mid-treatment clonality, and mid-treatment evenness were correlated with PET response status (Wilcoxon rank-sum p=0.031-0.048). None of the TCR diversity metrics were strongly correlated with mid-treatment residual total lesion glycolysis (median Spearman |r| = 0.17 [0.04-0.52]). Patients classified as mid-treatment PET responders had higher percentage of peripheral polyfunctional CD8+ T-cells compared to mid-treatment PET non-responders (Figure 5C, 0.7-1.3% vs. 0-0.1% expressing MIP1β and IFNγ). There was no association between total lesion glycolysis and peripheral CD8+ T-cell functionality.

**Figure 5.**
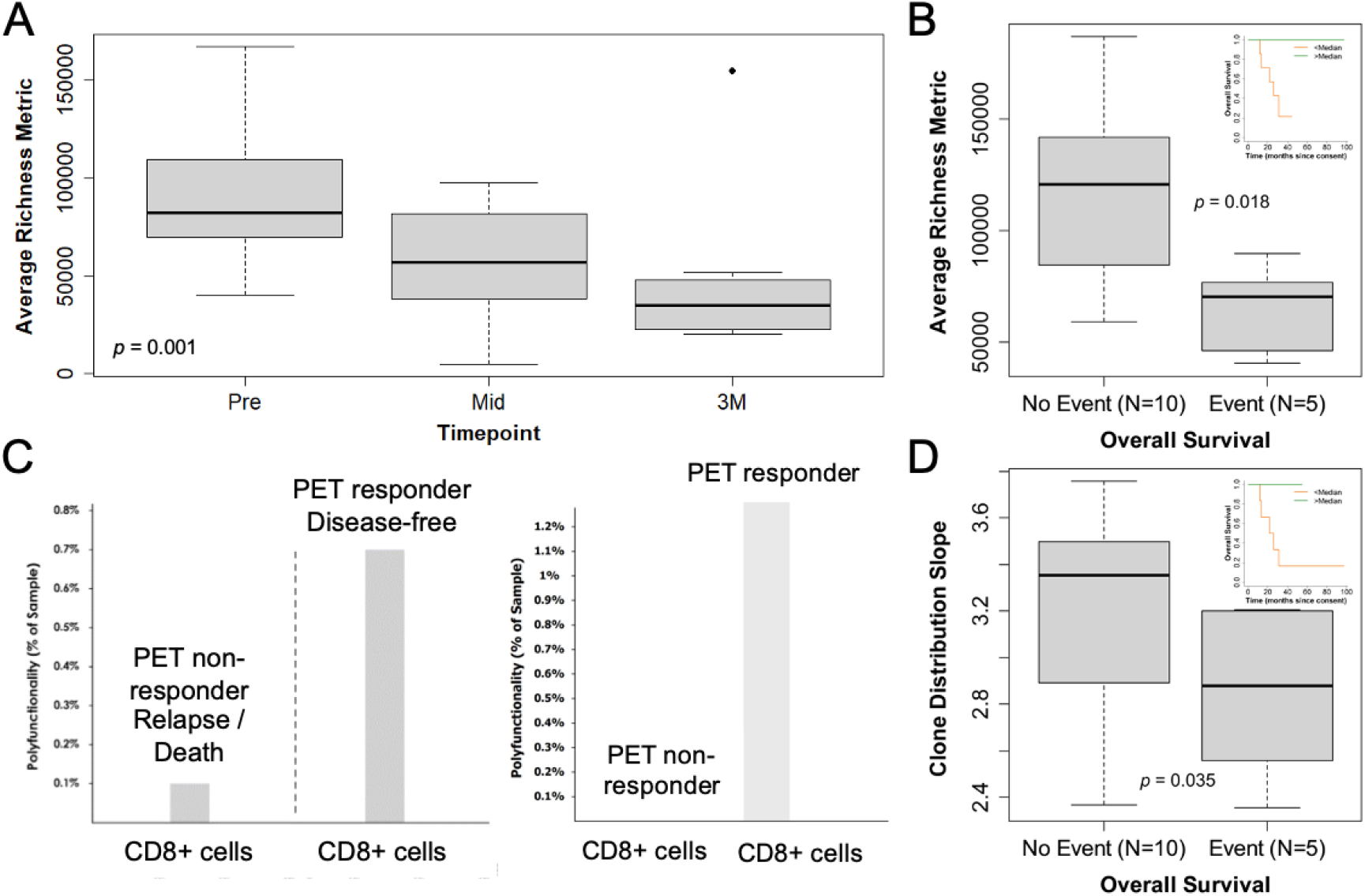
T-cell receptor (TCRβ CDR3) richness boxplots over pre/mid/post-treatment timepoints (A), TCR richness and clone distribution slope boxplots grouped by overall survival events with Kaplan-Meier insets (B,D), and CD8+ T-cell polyfunctionality for pairs of PET responders and PET non-responders (C).

### Plasma Cytokine Levels

Figure 6 displays the mid-treatment peripheral blood plasma cytokine hierarchical clustering dendrogram with heatmap scaled by z-scores of cytokine concentration levels and rugs for OS, PFS, and PET response status. While the 23 patients were not distinctly clustered by cytokine levels, hierarchical clusters of cytokines included the following: (i) MIP1β, IFNγ, TNFR1, TNFR2, TNFα; (ii) VEGF, TGFβ1. Among these select cytokines, lower mid-treatment TNFR1 relative to baseline was associated with worse PFS (HR 0.43, p=0.005) and higher mid-treatment TNFα relative to baseline was associated with worse OS (HR 1.59, p=0.05). Higher mid-treatment TGFβ1 showed a trend towards correlation with PET response (Fisher p=0.065).

**Figure 6.**
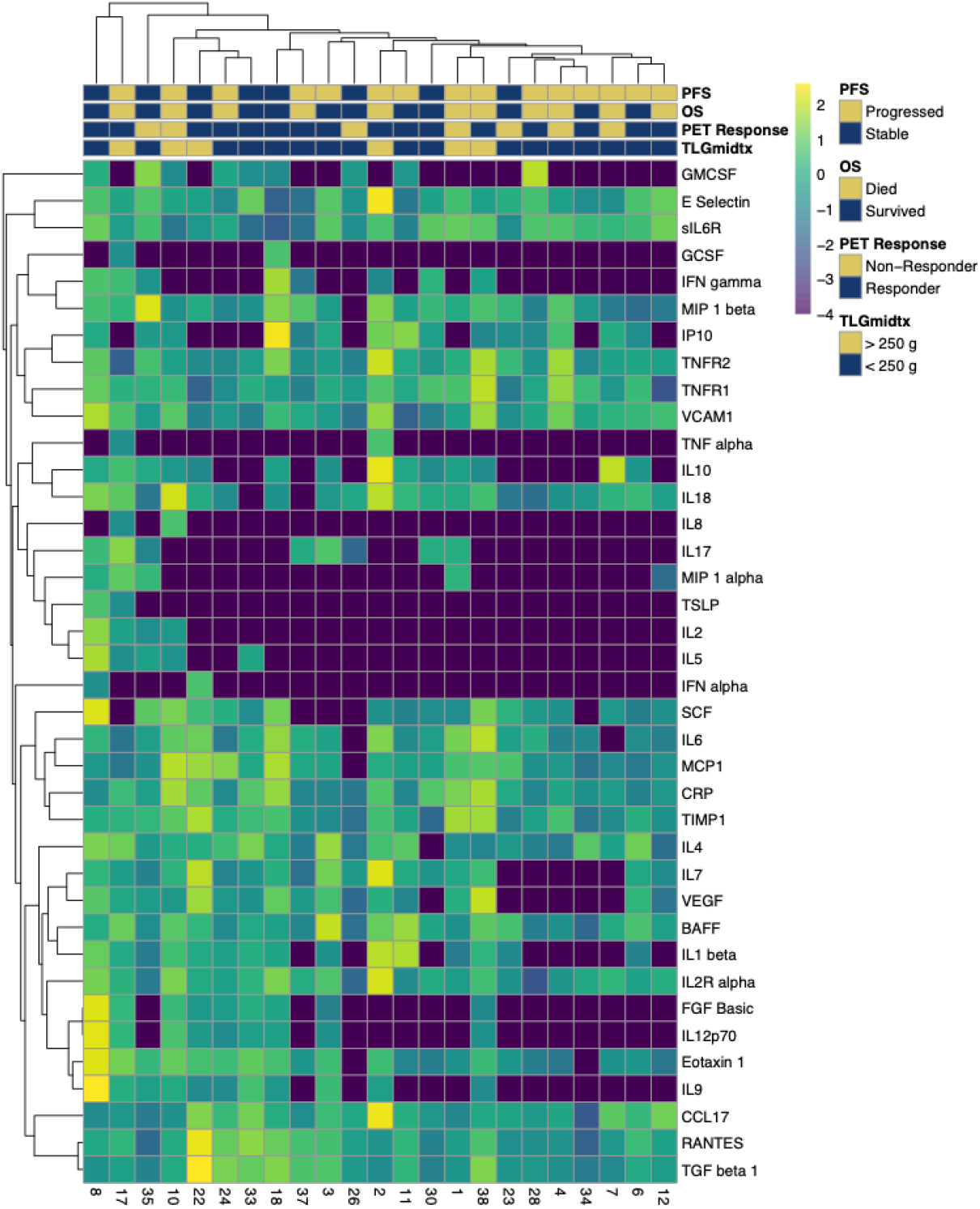
Mid-treatment peripheral blood plasma cytokines hierarchical cluster dendrogram heatmap with rugs for overall survival (OS), progression-free survival (PFS), and FLARE PET response status.

## Discussion

We conducted a phase II trial testing selective radiation dose escalation in patients classified as non-responders on mid-chemoradiation PET/CT (FLARE-RT trial), with the objective that this would improve overall survival due to better local control in patients at highest risk of local recurrence. All of the patients received concurrent platinum doublet chemotherapy, and approximately half the patients received durvalumab post-chemoradiation in accordance to the PACIFIC trial results that changed the standard of care for this population^2^. Although our trial population is not identical to the PACIFIC trial population (which only included patients who recovered from toxicity of chemoradiation) nor the patients in RTOG 0617 (none received durvalumab post-chemoradiation)^15^, our results compare favorably against both trials: our 1-year OS was 82% (vs 83.1% in PACIFIC and 80.0% in RTOG 0617), our 1-year PFS was 53% (vs 55.9% in PACIFIC and 49.2% in RTOG 0617), and our 1-year LRC was 88% (vs 83.7% in RTOG 0617, and 12.6% lung recurrence plus 6.5% nodal recurrence in PACIFIC). We saw an increase in both PFS and pneumonitis with durvalumab, although neither were statistically significant in our relatively small sample size. In our trial, disease recurrence was driven primarily by distant metastatic progression, with distant metastasis being a site of first recurrence in 19/23 (83%) patients. This suggests improvement in systemic disease control is needed in a subset of patients to further improve patient outcomes.

We found that mid-chemoradiation PET imaging independently predicts for OS and PFS. Multiple prior series have found changes in SUVmax during chemoradiation to be prognostic for survival ^9,43.^ In our trial, being a PET responder versus non-responder no longer conferred a difference in OS or PFS, perhaps due to the fact we dose-escalated patients based on PET changes during chemoradiation, mitigating the prognostic power of the PET response. However, similar to other published reports^4,9^,10, we did find that mid-treatment TLG independently predicted for OS and PFS, potentially serving as a marker of disease burden, even after adjustments for clinical and treatment factors. Patients classified as PET non-responders on the mid-treatment scan who also presented with a high residual TLG had the worst outcomes. This strategy could potentially help risk-stratify patients for further treatment intensification.

Our findings suggest a correlation between mid-treatment PET response to chemoradiation and peripheral immunologic status, although our results are only hypothesis generating and not conclusive. Mid-treatment PET response status was associated to PD-L1 expression, with PD-L1 positive tumors likely to be PET responders and PD-L1 negative tumors likely to be PET non-responders. Tumor response to radiation treatment requires an intact immune system with functional T-cells^22^, and radiation has been shown to induce a pro-inflammatory tumor microenvironment via the induction of an immunogenic cell death^44,45.^ Since PD-L1 expression plays a major role in suppressing adaptive immunity, it is possible that radiation treatment could produce a pro-inflammatory environment that helps overcome PD-L1 induced immune suppression.

Our analysis of germline DNA SNPs found differences in gene alteration frequency between PET responders and non-responder, particularly in immunology pathways, which also correlated with survival. The JAK-STAT signaling pathway plays critical roles in cytokine receptors and can modulate the polarization of T helper cells. A recent report by Shahamatdar et al.^46^ analyzing germline variants in the TCGA cohort demonstrated that host genetics are associated with phenotypes that describe the immune component of the tumor microenvironment; they found one SNP associated with the amount of infiltrating follicular helper T cells; and 23 candidate genes, some of which are involved in cytokine-mediated signaling. Our patient cohort supports prior findings that patients with greater TCR richness in the peripheral blood had improved survival compared with patients with less clonal richness^47-49^. TCR richness declined with chemoradiation although a smaller mid-treatment decline in TCR diversity was associated with improved survival. Pre-treatment TCR clone distribution slope, among other diversity metrics, was correlated with PET response status but none of the TCR diversity metrics were correlated with mid-treatment residual total lesion glycolysis. This suggests that PET-response status may be a marker of cancer/patient biology, whereas TLG may be an independent measure of cancer disease burden. Single cell functional assays were performed on the peripheral blood for a small subset of patients, suggesting mid-treatment PET responders had higher percentages of peripheral polyfunctional CD8+ T-cells compared to non-responders. We also saw that select plasma cytokines are associated with OS and PFS and correlated with PET response, including TNFα, TNFR1, TGFβ, and MIP1β.

In conclusion, in a prospective phase 2 trial of response-adaptive radiation dose escalation for patients with unresectable NSCLC, we confirm that mid-chemoradiation PET imaging is prognostic for survival and found that PET response status may be linked with peripheral T-cell function. The combination of PET response imaging and peripheral blood biomarkers could be used to guide further clinical trials of treatment intensification by identifying patients at highest risk of treatment failure.

## Data Availability

Data will be made publicly available and upon request at the conclusion of the clinical trial.

## Acknowledgments

This work was funded by NIH / NCI R01CA204301. We thank Priya Vissamraju and Christina Lo for coordination of the FLARE-RT clinical trial. We acknowledge the efforts of nuclear medicine and radiation oncology technologists during PET/CT acquisitions, radiation therapy planning, and image-guided radiation therapy delivery.

---

**Supplemental Table 1.**
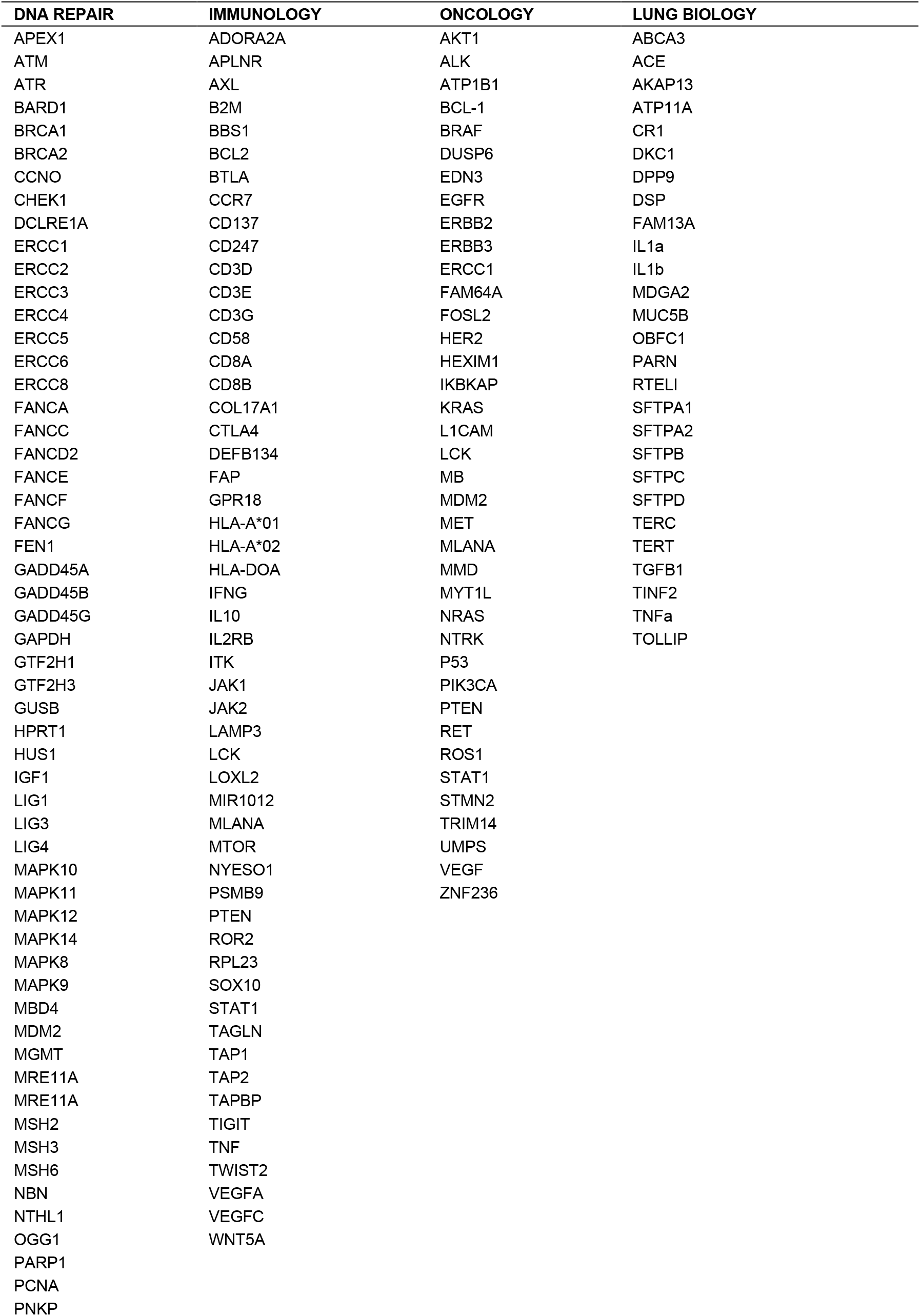
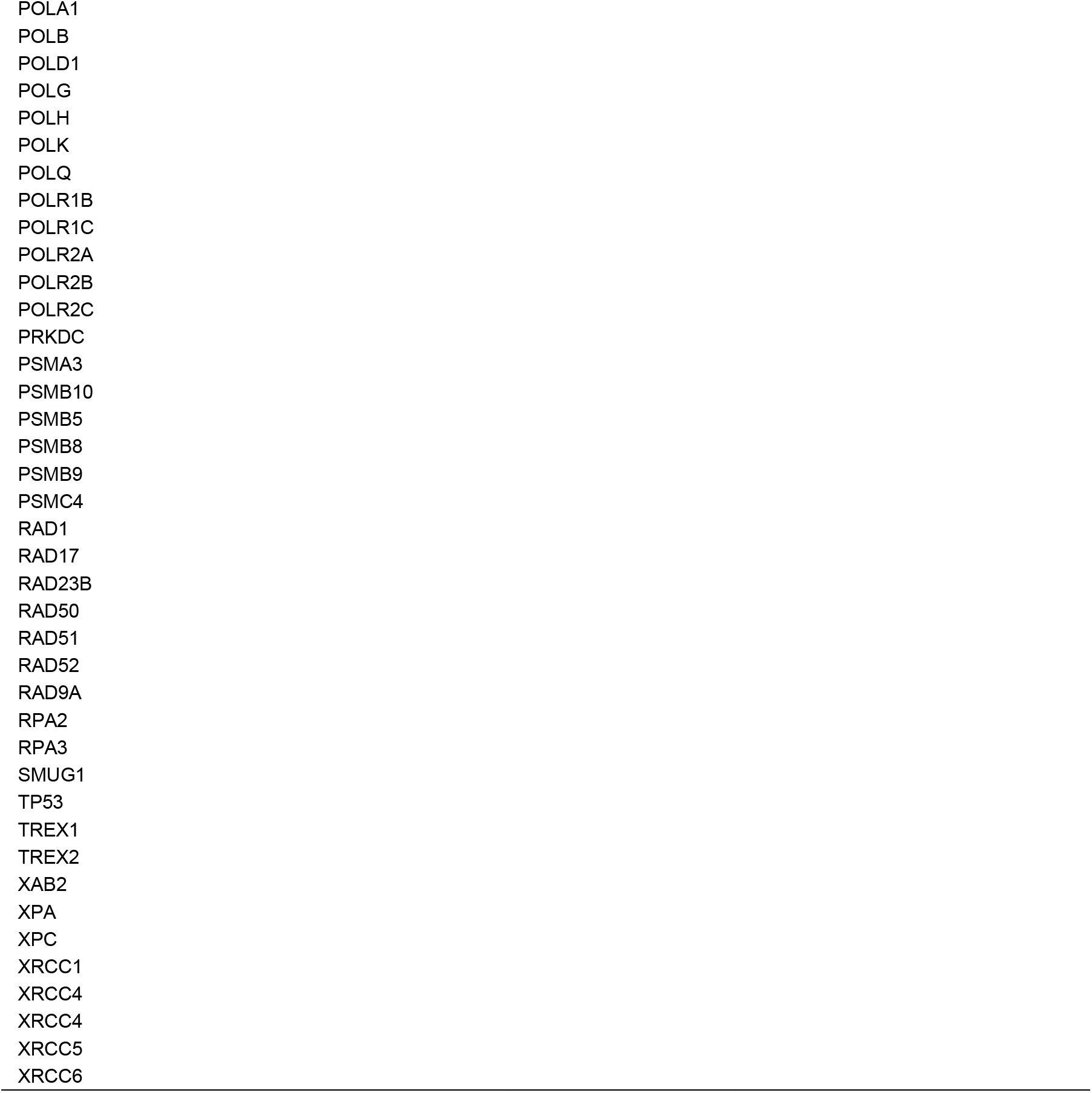
Predefined Candidate Genes for 4 Pathways: DNA Repair, Immunology, Oncology, and Lung Biology

